# Age-Related Speech-in-Noise Hearing Loss in Parkinson’s Disease and *APOE* E4 Carriers

**DOI:** 10.64898/2026.06.08.26355175

**Authors:** Matthew J. Kmiecik, Wanwan Xu, Catherine H. Weldon, Anna Guan, Matthew H. McIntyre, Ellen L. Bouchard, 23andMe Research Team, Ruth B. Schneider, Adam Auton, Stella Aslibekyan

## Abstract

Age-related hearing loss is a leading modifiable risk factor for dementia and is increasingly recognized as a non-motor feature of Parkinson’s disease (PD). The apolipoprotein E (*APOE*) E4 allele is the strongest genetic risk factor for Alzheimer’s disease and is associated with cognitive decline in PD, yet its relationship to hearing loss remains unclear. Therefore, we examined the independent and interactive effects of PD status and *APOE* E4 carrier status on age-related hearing loss using a validated web-based speech-in-noise (SIN) assessment in 239,620 23andMe Research Institute participants without PD and 4,361 PD cases. Generalized additive models for location, scale, and shape (GAMLSS) showed that both PD and *APOE* E4 independently exacerbated age-related hearing decline, with speech reception thresholds (SRTs) worsening non-linearly with advancing age, but without evidence of synergistic interaction. However, longitudinal analyses in a subcohort completing at least two assessments (1,434 PD cases; 36,242 controls) using GAMLSS mixed models showed a significant three-way interaction between PD status, *APOE* E4, and age^2^, such that SIN hearing loss accelerated more steeply with age in *APOE* E4 carriers with PD. Males and individuals with lower educational attainment also exhibited worse SIN hearing loss. These results identify *APOE* E4 carriers with PD as a priority population for hearing screening and intervention, and support the integration of SIN assessments into routine PD care to detect hearing decline that may compound cognitive and communicative burden in aging.

## Main

Many people experience the gradual effects of age-related hearing loss in later life, with clinically significant hearing loss impacting more than two thirds of all adults aged 60 years or older (Lin, 2024). Age-related hearing loss is a risk factor for incident cognitive impairment and dementia, including Alzheimer’s disease (AD) (Yu et al., 2024). Hearing loss is also associated with increased risk and incidence of Parkinson’s disease (PD) (Lai et al., 2014; Readman et al., 2025; Simonet et al., 2022) and several studies have demonstrated worse hearing ability in individuals with PD relative to matched controls (Jafari et al., 2020; Rajai Firouzabadi et al., 2026), leading many to suggest that hearing loss may present as a prodromal non-motor feature of PD. While recent Mendelian randomization studies have found no causal relationship between hearing loss and PD (Ning et al., 2024; Zhang et al., 2025), the clear association between these two conditions from several observational studies suggest a shared underlying pathology that may explain their co-occurrence. Additionally, these Mendelian randomization approaches were limited in their ability to characterize the nature of auditory dysfunction, and further work using more precise phenotypic measures is needed to fully understand the relationship between hearing and PD.

Hearing loss is one of the largest modifiable risk factors for dementia through the use of hearing-protective devices, such as earplugs, and hearing-assistive devices, such as hearing aids and cochlear implants (Livingston et al., 2020). Indeed, dispensation of hearing aids was shown to decrease the incidence of PD in an observational study (Neilson et al., 2024). Secondary analyses of the Aging and Cognitive Health Evaluation in Elders (ACHIEVE) study—a 3-year randomized hearing intervention in older adults with untreated hearing loss and without cognitive impairment—demonstrated that hearing aid dispensation slowed cognitive decline most in individuals with greater risk of dementia (Lin et al., 2023; Pike et al., 2025). Although randomized controlled clinical trials of hearing interventions and PD risk are absent, the results from ACHIEVE and observational studies show promise of hearing loss as a modifiable risk factor for PD, especially in individuals at greatest risk for cognitive impairment.

*APOE* E4 is the strongest genetic risk factor for AD dementia and cognitive decline (Kloske et al., 2024; Narasimhan et al., 2024). Likewise, *APOE* E4 carrier status in individuals with PD is associated with worse cognitive outcomes, faster cognitive decline, and increased risk of PD dementia (Kmiecik et al., 2025; Rosal et al., 2025; Szwedo et al., 2022). The evidence for an association between *APOE* E4 and hearing impairment, however, is mixed for both peripheral pathways measured via pure-tone audiometry (Cha et al., 2024; Han et al., 2024) and more centrally-mediated auditory processing (Jayakody et al., 2020; Tuwaig et al., 2017; Zimmermann et al., 2019). Given that the effects of *APOE* E4 on hearing loss may be subtle and heterogeneous, previous studies may have been underpowered, especially to detect interactions between genotype and age (Han et al., 2024).

Therefore, we examined the relationship between *APOE* E4 and PD on age-related hearing loss using a validated web-based adaptive speech-in-noise (SIN) hearing assessment in the 23andMe Research Institute cohort. Because difficulty understanding SIN is the most commonly reported auditory complaint among individuals with hearing loss (Billings et al., 2023), SIN performance offers a clinically and ecologically valid index of real-world hearing function. PD is associated with hearing loss across both peripheral and central pathways (Jafari et al., 2020; Rajai Firouzabadi et al., 2026), and *APOE* E4 is mainly associated with cognitive decline; therefore, we hypothesized that *APOE* E4 and PD would interact with age to exacerbate age-related hearing loss in carriers and cases of PD.

## Methods

### Participants

23andMe Research Institute research participants between 50–85 years of age were recruited to take a validated web-based hearing test with accompanying self-report survey every 6 months for 3 years. Participants were able to access the hearing survey and test via the 23andMe web interface and email reminders were sent to participants that began but did not complete the survey. Participants were not able to complete the hearing test on their mobile phones or devices via the 23andMe application and were instead instructed to complete the hearing test via a web browser on a laptop or desktop computer. Participants provided informed consent and volunteered to participate in the research online, under a protocol approved by the external AAHRPP-accredited Salus IRB (https://www.versiticlinicaltrials.org/salusirb). Accordingly, only participants from the United States and United Kingdom were included in analyses given legal/regulatory compliance policies. All data were obtained from a 23andMe Research Institute database cache dated 03 April 2026.

### Genetic Data

Saliva-based genotyping was performed on all participants across five 23andMe genotyping platforms (see Supplementary Materials). Two single nucleotide polymorphisms (SNPs; rs429358 and rs7412) were used to determine the number of *APOE* E4 alleles. Participants were classified as *APOE* E4 carriers (one or two E4 alleles) or non-carriers (zero E4 alleles). Participants carrying the *LRRK2* p.G2019S or *GBA1* p.N409S (previously N370S) variants were excluded to focus on idiopathic PD cases and reduce genetic confounding. Following the methods of established 23andMe genome-wide association study (GWAS) practices (23andMe Research Team, 2024), related participants sharing more than 700 centimorgan of identical by descent DNA were excluded, preferentially retaining PD cases. Genetic ancestry groups (23andMe, Inc., 2022; Durand et al., 2014) and principal components (PCs) were computed from genome-wide SNP data to account for population stratification (see Supplementary Materials). Participants’ sex (female/male) was determined via genotyping.

### Hearing Test

Participants first completed a self-report survey that queried participants’ auditory symptoms, hearing device usage, previous hearing-related medical and family history, and hearing loss characteristics. Participants were also asked two questions: whether they were fluent in English (yes/no) and whether they were using headphones (in-ear or over-hear) or speakers to complete the hearing assessment. Following the survey, participants completed a validated web-based SIN hearing assessment developed by hearX^®^ (De Sousa et al., 2018; Potgieter et al., 2016). Participants were tasked with identifying spoken digit triplets (0–9) amongst background noise, of which the signal-to-noise ratio (SNR) was modulated by 2 decibels (dB) according to participants’ accuracy on the previous trial (lower and higher SNR for correct and incorrect response, respectively; see Figure 1). Participants entered the digits they heard using the keyboard or mouse click. A total of 23 trials were presented to participants and the first four trials were excluded to allow the test to adapt to participants’ hearing ability. An anticipated SNR value was generated for trial 24 that was never presented. Speech reception thresholds (SRT) were quantified by averaging the SNR across trials 5–23 and the generated SNR for trial 24 for a total of 19 SNR values in dB. Lower (more negative) SRT values indicate better SIN hearing ability, while higher values indicate worse SIN hearing performance.

**Figure 1.**
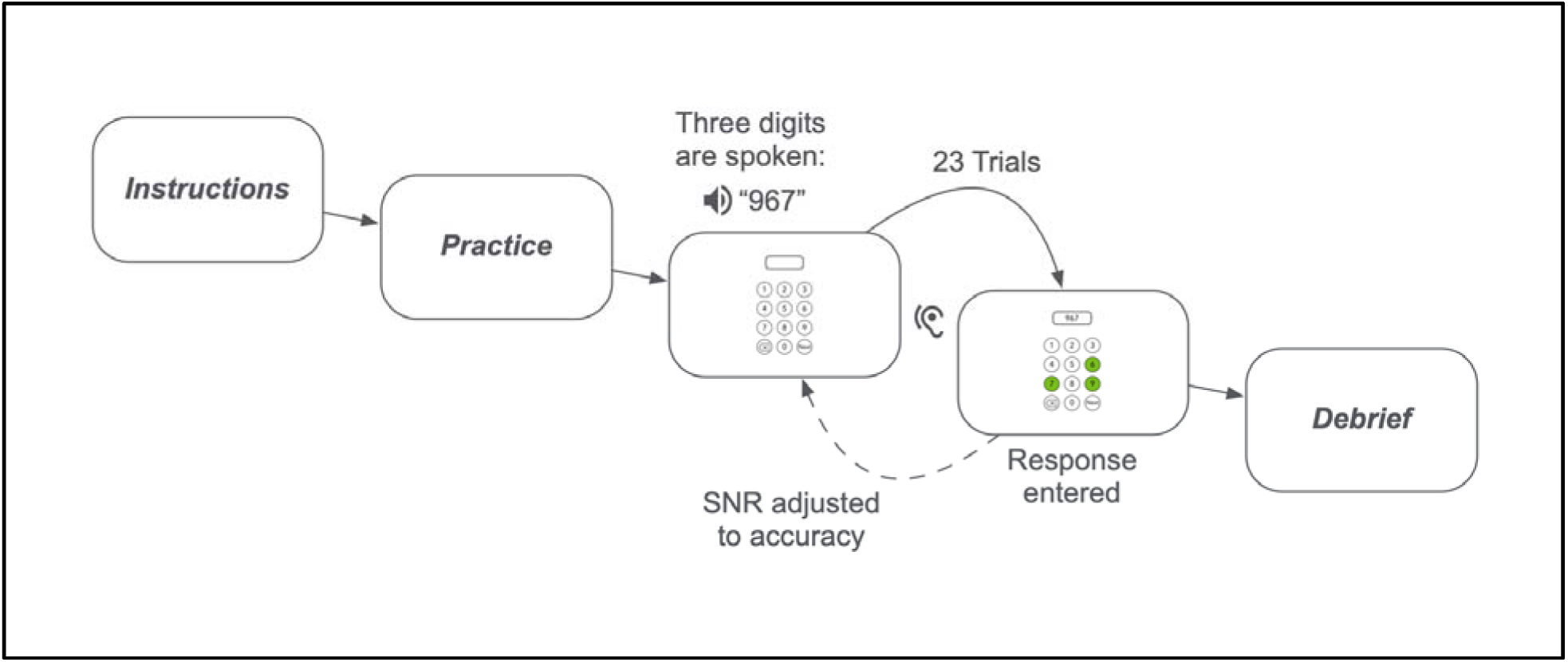
Hearing assessment trial paradigm. After giving instructions and practice, participants listened to 23 trials of spoken digit triplets in English amongst background noise and entered responses using the keyboard or mouse. The signal-to-noise ratio (SNR) of the subsequent trial is reduced for correctly answered trials and increased for incorrect trials. Participants were debriefed with their hearing ability and resources to learn more.

### Parkinson’s Disease Status and Additional Participant Characteristics

Additional participant information was collected from other cross-sectional and longitudinal surveys available to 23andMe Research Institute participants. Participants’ education level was binarized into less than Associate degree and Associate degree or greater. Participants’ PD case/control status was determined based on self-reported diagnosis. To be considered controls, participants must have reported “no” to a diagnosis of PD. Participants that initially reported a PD diagnosis but later revised their diagnosis status were excluded. Additional PD characteristics like age of PD diagnosis, if available, were aggregated for descriptive purposes. We derived case-control status from the cumulative self-reported PD status, and SRT measurement may have preceded or followed PD diagnosis. Consistent with previous work investigating SRTs in PD, we did not adjust for hearing aid usage (Readman et al., 2025).

## Statistical Analysis

Participants with SRT values outside the 99^th^ percentile (i.e., in the bottom 0.5% and top 0.5% of the SRT distribution) were identified as outliers and excluded (see Figure S1). We further excluded a small fraction of participants who were not fluent in English and reported using a device other than headphones or speakers. Given attrition following baseline assessments in longitudinal designs, we analyzed the data using two cohorts of participants: participants that completed 1) a baseline hearing assessment (i.e., cross-sectional) and 2) at least two hearing assessments (i.e., longitudinal). In both baseline and longitudinal models, we estimated SRT as a function of PD status, *APOE* E4 carrier status, and their factorial interactions with age and age^2^. Covariates included sex, education, speaker device, genotyping platform, and 10 ancestry PCs. Age was mean centered. To balance PD cases and controls, models were weighted by coarsened exact matching (CEM) weights based on participants’ age (in five-year bins), sex, and education. CEM weights were estimated separately for each participant cohort, reducing the effective sample size (ESS) of controls from *N*=239,620 to ESS=170,103 in the baseline cohort and from *N*=36,242 to ESS=29,467 in the longitudinal cohort; PD cases were retained at full weight in both cohorts.

We initially explored using a linear regression approach to modeling SRT data. However, our observed SRT distribution was positively skewed (see Figure S1) resulting in non-normally distributed residuals (see Figure S2). Therefore, to increase model validity and inferential ability, we employed generalized additive models for location, scale, and shape (GAMLSS) (Rigby & Stasinopoulos, 2005) and compared the model diagnostics from both skew-normal and skew-t distributions (see Figure S2, Table S1). On the basis of normally distributed residuals, well-fitting QQ-plots, and significantly lower Akaike Information Criterion (AIC) and Bayesian Information Criterion (BIC) values, the GAMLSS model with a skew-t distributional family was best suited for our SRT outcome and was utilized for both baseline and longitudinal cohort analyses. A GAMLSS mixed-model estimated longitudinal change in SRT as a function of the predictors outlined above with the addition of an effect of time since baseline in years and a random effect of intercept for participants. We used participants’ mean-centered baseline age to reduce collinearity with time. To improve convergence of the four-parameter skew-t distribution, we adopted a staged warm-start approach in which ν (skewness parameter) and τ (tail weight) were fixed at values estimated from a preliminary simpler model of baseline SRT performance from the longitudinal cohort. Sensitivity analyses excluding non-Europeans (i.e., individuals not classified as genetically European) did not change results.

Data processing was performed in python v.3.12.5 (Python Software Foundation, 2016) and R v.4.4.0 (R Core Team, 2024) using the following packages: *pandas* (The pandas development, 2026), *numpy* (Harris et al., 2020), and *tidyverse* (Wickham et al., 2019) for data wrangling and processing, *gamlss* (Stasinopoulos & Rigby, 2008) for GAMLSS modeling*, marginaleffects* (Arel-Bundock et al., 2024) for average marginal effects and predictions, and *datawizard* (Patil et al., 2022) for variable standardization.

## Results

### Baseline Age-Related Hearing Loss

After excluding *n*=10,946 genetically related participants, a total of *n*=243,981 participants were eligible for analysis of their baseline SRT: *n*=4,361 PD cases (23.27% *APOE* E4 carriers) and *n*=239,620 controls (25.55% *APOE* E4 carriers). See Table 1 for participant demographics and characteristics weighted by CEM (see Table S2 for an unweighted table). Within PD cases, the age of PD diagnosis was similar between *APOE* E4 carriers and non-carriers.

**Table 1.** Participant demographics and characteristics at baseline for the cross-sectional cohort weighted by coarsened exact matching.

| Characteristic | Sub-Category | Parkinson's Disease (PD) Status |  |  |  |
| --- | --- | --- | --- | --- | --- |
|  |  | Control |  | PD |  |
|  |  | E4- | E4+ | E4- | E4+ |
| N |  | 178,402 | 61,218 | 3,346 | 1,015 |
| Age |  | 69.3 (0.021) | 68.81 (0.035) | 69.51 (0.126) | 68.58 (0.224) |
| Sex | Male | 58.8% (0.1%) | 58.9% (0.2%) | 59.1% (0.8%) | 57.9% (1.5%) |
| Education | ≥ Assoc. degree | 76.3% (0.1%) | 76.8% (0.2%) | 75.7% (0.7%) | 78.8% (1.3%) |
| Genetic Ancestry | European | 87.6% (0.1%) | 89.6% (0.1%) | 91.4% (0.5%) | 92.5% (0.8%) |
| Age at PD diagnosis |  | - | - | 63.52 (0.172) | 63.23 (0.295) |
| APOE E4 status | Homozygous | - | 7% (0.1%) | - | 7.4% (0.8%) |
| Speech reception threshold (dB) |  | -7.18 (0.008) | -7.22 (0.014) | -6.78 (0.051) | -6.78 (0.099) |
| Device used for test | Headphones | 24.1% (0.1%) | 25.2% (0.2%) | 22.7% (0.7%) | 26.9% (1.4%) |
| Background noise level (0-5) |  | 0.95 (0.003) | 0.95 (0.006) | 1.01 (0.02) | 0.99 (0.037) |
| Difficulty hearing in noise | A lot of difficulty | 15.8% (0.1%) | 15.7% (0.2%) | 18% (0.7%) | 17.6% (1.2%) |
| Self-reported hearing problems | Yes | 44.4% (0.1%) | 44% (0.3%) | 44% (0.9%) | 43.1% (1.6%) |
| Degree of hearing loss | None/Mild | 54% (0.2%) | 54% (0.4%) | 52.5% (1.3%) | 55.4% (2.5%) |
| Diagnosed with hearing loss | Yes | 93.7% (0.1%) | 93.2% (0.2%) | 94.2% (0.7%) | 93.2% (1.4%) |
| Age at hearing loss diagnosis |  | 55.91 (0.092) | 55.04 (0.162) | 55.02 (0.514) | 54.77 (0.974) |
| Use cochlear implant | Yes | 0.1% (0%) | 0.1% (0%) | 0.2% (0.1%) | Masked |
| Age started cochlear implant |  | 62.02 (0.996) | 59.93 (1.941) | 67.86 (2.781) | Masked |
| Use hearing aid | Yes | 16.8% (0.1%) | 16.1% (0.2%) | 18.1% (0.7%) | 17.4% (1.2%) |
| Age started hearing aid |  | 64.68 (0.081) | 64.18 (0.144) | 64.07 (0.432) | 63.36 (0.863) |
| Use assistive listening device | Yes | 0.6% (0%) | 0.6% (0%) | 0.7% (0.1%) | 1% (0.3%) |
| Age started assistive device |  | 63.44 (0.432) | 63.76 (0.742) | 64.5 (1.75) | 59.64 (2.086) |
| No hearing devices | Yes | 82.8% (0.1%) | 83.4% (0.2%) | 81.4% (0.7%) | 81.9% (1.2%) |
| Ménière's disease | Yes | 2.1% (0%) | 2.1% (0.1%) | 2.1% (0.3%) | 2.7% (0.5%) |
| Vertigo | Yes | 35.2% (0.1%) | 35% (0.2%) | 41.2% (0.9%) | 40.2% (1.6%) |
| Tinnitus | Yes | 37.1% (0.1%) | 37.5% (0.2%) | 37.5% (0.9%) | 34.3% (1.5%) |
| Tinnitus severity (0-10) |  | 2.57 (0.013) | 2.6 (0.022) | 2.7 (0.072) | 2.65 (0.138) |

Note. Categorical measures presented as % (SE), continuous as Mean (SEM). N reflects unweighted participant counts; all statistics (means, percentages) are weighted by coarsened exact matching (CEM). CEM weights reduced the effective sample size (ESS) of controls from $N=243,981$ to $ESS=170,103$ (cases retained at full weight). E4+ indicates APOE E4 carriers, E4- non-carriers. Indented items are contingent follow-ups. Survey response percentages calculated among respondents providing definitive answers; non-responders and "I'm not sure" responses excluded from denominators. Statistics with $n<5$ masked for privacy; dashes indicate not applicable. SRT=speech reception threshold.

The GAMLSS model in the cross-sectional baseline cohort demonstrated that males had more hearing loss than females and that greater levels of education were associated with better hearing ability, over and above covariates that included ancestry PCs, genotyping platform, and speaker device (see Table 2). Hearing loss increased nonlinearly with age, and was greater in PD cases relative to controls, and greater in *APOE* E4 carriers relative to non-carriers (see Figure 2). However, we did not observe any interactions between age, PD status, or *APOE* E4 in the baseline cohort model.

**Table 2.** Baseline GAMLSS Model Results.

| Term | $b$ | 95% CI | | SE | $t$ | $p$ | $-\log_{10}(p)$ |
| --- | --- | --- | --- | --- | --- | --- | --- |
|  |  | Lower | Upper |  |  |  |  |
| Intercept | -10.009 | -10.029 | -9.988 | 0.010 | -965.72 | < mp | 202517.58 |
| Sex (Male) | 0.342 | 0.328 | 0.356 | 0.007 | 49.14 | < mp | 526.14 |
| Edu. (≥Assoc.) | -0.209 | -0.224 | -0.193 | 0.008 | -26.04 | 1.60e-149 | 148.80 |
| Age | 0.078 | 0.077 | 0.079 | 0.001 | 119.90 | < mp | 3124.09 |
| Age<sup>2</sup> | 0.002 | 0.002 | 0.002 | 5.93e-05 | 32.70 | 1.58e-234 | 233.80 |
| PD (Case) | 0.252 | 0.176 | 0.329 | 0.039 | 6.45 | 1.14e-10 | 9.94 |
| E4 (Carrier) | 0.030 | 0.010 | 0.051 | 0.010 | 2.94 | 0.003 | 2.48 |
| PD × Age | 3.07e-04 | -0.010 | 0.010 | 0.005 | 0.06 | 0.951 | 0.02 |
| PD × Age<sup>2</sup> | -2.94e-04 | -0.001 | 0.001 | 4.62e-04 | -0.64 | 0.525 | 0.28 |
| E4 × Age | 0.002 | -5.62e-05 | 0.005 | 0.001 | 1.92 | 0.055 | 1.26 |
| E4 × Age<sup>2</sup> | 5.39e-05 | -1.80e-04 | 2.88e-04 | 1.19e-04 | 0.45 | 0.652 | 0.19 |
| PD × E4 | -0.046 | -0.203 | 0.111 | 0.080 | -0.58 | 0.564 | 0.25 |
| PD × E4 × Age | 0.001 | -0.019 | 0.020 | 0.010 | 0.07 | 0.948 | 0.02 |
| PD × E4 × Age<sup>2</sup> | 0.001 | -0.001 | 0.003 | 0.001 | 1.16 | 0.248 | 0.61 |
| PC0 | 0.013 | 0.005 | 0.021 | 0.004 | 3.30 | 0.001 | 3.01 |
| PC1 | -0.031 | -0.039 | -0.024 | 0.004 | -8.03 | 9.61e-16 | 15.02 |
| PC2 | -0.049 | -0.057 | -0.042 | 0.004 | -12.56 | 3.44e-36 | 35.46 |
| PC3 | 0.004 | -0.003 | 0.011 | 0.004 | 1.20 | 0.232 | 0.63 |
| PC4 | -0.030 | -0.038 | -0.022 | 0.004 | -7.55 | 4.23e-14 | 13.37 |
| PC5 | -0.014 | -0.021 | -0.006 | 0.004 | -3.60 | 3.13e-04 | 3.51 |
| PC6 | 0.004 | -0.004 | 0.012 | 0.004 | 1.02 | 0.306 | 0.51 |
| PC7 | -0.017 | -0.023 | -0.010 | 0.003 | -5.03 | 4.89e-07 | 6.31 |
| PC8 | -0.004 | -0.011 | 0.004 | 0.004 | -0.99 | 0.324 | 0.49 |
| PC9 | 0.022 | 0.015 | 0.030 | 0.004 | 6.20 | 5.66e-10 | 9.25 |
| Platform V2 | 0.047 | -0.057 | 0.152 | 0.053 | 0.88 | 0.377 | 0.42 |
| Platform V3 | 0.029 | -0.008 | 0.067 | 0.019 | 1.52 | 0.129 | 0.89 |
| Platform V4 | -0.012 | -0.031 | 0.006 | 0.010 | -1.30 | 0.192 | 0.72 |
| Device: Headphones | -0.883 | -0.899 | -0.868 | 0.008 | -112.74 | < mp | 2762.14 |

Note. < mp denotes $p$ -values smaller than the machine precision (see $-\log_{10}(p)$ for more precise $p$ -values); E4 denotes APOE E4 carrier status; PD=Parkinson's disease status; PC=ancestry principal component.

**Figure 2.**
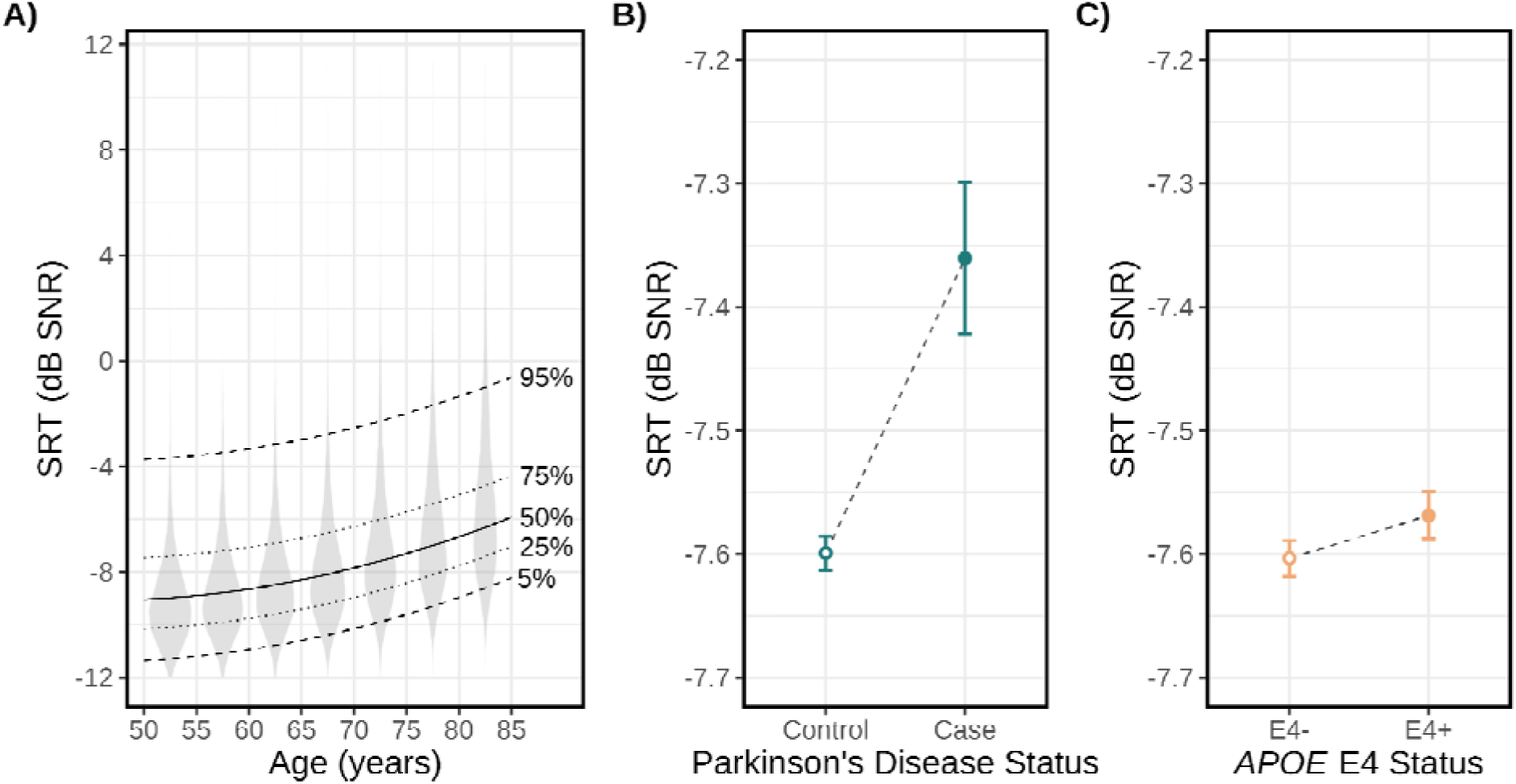
Hearing loss accelerates with age, is greater in Parkinson’s Disease (PD) cases relative to controls, and greater in *APOE* E4 carriers relative to non-carriers. Greater speech reception thresholds (SRTs) indicate increased hearing loss. A) Predicted percentile curves (5th, 25th, 50th, 75th, and 95th) of SRT across age, derived from the fitted skew-t distribution using age-varying average marginal predictions from 5,000 randomly sampled participants. Observed data are represented by violin plots. B) Average marginal effects (AMEs) of PD on SRTs. C) AMEs of *APOE* E4 on SRTs. All effects were estimated from the cross-sectional GAMLSS skew-t model at participants’ baseline hearing assessment. Y-axes in B) and C) are restricted to the range of the estimates to highlight effect magnitude. *APOE* E4 status is designated as E4+ for carriers and E4- for non-carriers; dB=decibel; SNR=signal-to-noise ratio. Error bars are 95% CIs.

### Longitudinal Changes in Hearing Loss

After excluding 382 genetically related participants, a total of *n*=37,676 participants completed at least two SIN hearing tests and were eligible for longitudinal analysis: *n*=1,434 PD cases (21.13% *APOE* E4 carriers) and *n*=36,242 controls (25.53% *APOE* E4 carriers). See Table 3 for participant demographics and characteristics weighted by CEM (see Table S3 for an unweighted table). A total of *n*=12,184 (32.34%) participants completed ≥3 observations, *n*=4,365 (11.59%) ≥4, *n*=1,721 (4.57%) ≥5, and *n*=658 (1.75%) all six. The mean time-interval between observations was 0.89 years (*SD*=0.56).

**Table 3.** Participant demographics and characteristics at baseline for the longitudinal cohort weighted by coarsened exact matching.

| Characteristic | Sub-Category | Parkinson's Disease (PD) Status |  |  |  |
| --- | --- | --- | --- | --- | --- |
|  |  | Control |  | PD |  |
|  |  | E4- | E4+ | E4- | E4+ |
| N |  | 26,990 | 9,252 | 1,131 | 303 |
| Age |  | 70.01 (0.046) | 69.56 (0.077) | 70.16 (0.198) | 69.43 (0.386) |
| Sex | Male | 59.1% (0.3%) | 58.9% (0.6%) | 59.2% (1.5%) | 58.4% (2.8%) |
| Education | ≥ Assoc. degree | 78.5% (0.3%) | 79.2% (0.5%) | 77.3% (1.2%) | 83.8% (2.1%) |
| Genetic Ancestry | European | 89.3% (0.2%) | 91.5% (0.3%) | 91.1% (0.8%) | 94.7% (1.3%) |
| Age at PD diagnosis |  | - | - | 64.77 (0.269) | 64.65 (0.507) |
| APOE E4 status | Homozygous | - | 7.1% (0.3%) | - | 5.9% (1.4%) |
| Speech reception threshold (dB) |  | -7.2 (0.019) | -7.22 (0.033) | -6.8 (0.084) | -6.81 (0.176) |
| Device used for test | Speaker | 74.5% (0.3%) | 73.5% (0.5%) | 77.1% (1.2%) | 71.3% (2.6%) |
| Background noise level (0-5) |  | 0.89 (0.007) | 0.9 (0.012) | 0.99 (0.033) | 0.94 (0.069) |
| Difficulty hearing in noise | A lot of difficulty | 17.3% (0.3%) | 17% (0.4%) | 17.8% (1.1%) | 18.3% (2.2%) |
| Self-reported hearing problems | Yes | 46.5% (0.4%) | 46.3% (0.6%) | 44.6% (1.5%) | 44% (3%) |
| Degree of hearing loss | None/Mild | 58.1% (0.5%) | 59.5% (0.8%) | 56.5% (2.1%) | 59.9% (4.1%) |
| Diagnosed with hearing loss | Yes | 92.4% (0.3%) | 91.3% (0.5%) | 92.1% (1.3%) | 89.5% (2.8%) |
| Age at hearing loss diagnosis |  | 56.57 (0.192) | 55.92 (0.339) | 56.21 (0.816) | 57.74 (1.674) |
| Use cochlear implant | Yes | 0.2% (0%) | 0.1% (0%) | Masked | - |
| Age started cochlear implant |  | 61.24 (1.862) | 65.52 (2.938) | Masked | - |
| Use hearing aid | Yes | 17.8% (0.3%) | 16.9% (0.4%) | 18.2% (1.1%) | 18.9% (2.3%) |
| Age started hearing aid |  | 65.51 (0.161) | 65.31 (0.269) | 65.79 (0.585) | 67.17 (1.065) |
| Use assistive listening device | Yes | 0.7% (0.1%) | 0.6% (0.1%) | Masked | Masked |
| Age started assistive device |  | 63.49 (0.686) | 64.99 (1.097) | 63.57 (2.885) | 64.86 (2.453) |
| No hearing devices | Yes | 81.7% (0.3%) | 82.6% (0.4%) | 81.5% (1.2%) | 80.5% (2.3%) |
| Ménière's disease | Yes | 2.2% (0.1%) | 2.1% (0.2%) | 2.7% (0.5%) | Masked |
| Vertigo | Yes | 37.1% (0.3%) | 36.7% (0.6%) | 40.4% (1.5%) | 44.3% (2.9%) |
| Tinnitus | Yes | 39.7% (0.3%) | 40.5% (0.6%) | 40% (1.5%) | 34% (2.8%) |
| Tinnitus severity (0-10) |  | 2.4 (0.027) | 2.41 (0.044) | 2.35 (0.107) | 2.33 (0.229) |

*Note.* Categorical measures presented as % (SE), continuous as Mean (SEM). N reflects unweighted participant counts; all statistics (means, percentages) are weighted by coarsened exact matching (CEM). CEM weights reduced the effective sample size (ESS) of controls from N=36,242 to ESS=29,467.1 (cases retained at full weight). E4+ indicates APOE E4 carriers, E4- non-carriers. Indented items are contingent follow-ups. Survey response percentages calculated among respondents providing definitive answers; non-responders and "I'm not sure" responses excluded from denominators. Statistics with $n < 5$ masked for privacy; dashes indicate not applicable. SRT=speech reception threshold.

Consistent with the baseline cohort model, the GAMLSS mixed model observed that SIN hearing loss was greater in males and participants with less education relative to females and greater education levels, respectively (see Table 4). We observed an effect of time (years since baseline) on SRTs, suggesting that, on average, SIN hearing loss worsened in our cohort within the allowed maximum of 3 years of repeated testing. SIN hearing loss increased nonlinearly with age, and interacted with PD status and *APOE* E4 carrier status. Importantly, a three-way interaction between these predictors was observed (*p*=1.71e-04). The positive PD × E4 × age^2^ coefficient indicates that SIN hearing loss accelerates more steeply with age in *APOE* E4 carriers with PD relative to all other groups with differences beginning to emerge at 70 years of age, especially in participants with PD (see Figure 3).

**Table 4.** Longitudinal GAMLSS Mixed Model Results.

| Term | b | 95% CI |  | SE | t | p | -log<sub>10</sub>(p) |
| --- | --- | --- | --- | --- | --- | --- | --- |
|  |  | Lower | Upper |  |  |  |  |
| Intercept | -8.908 | -8.921 | -8.895 | 0.007 | -1334.42 | < mp | 386674.97 |
| Sex (Male) | 0.511 | 0.502 | 0.520 | 0.005 | 113.24 | < mp | 2786.51 |
| Edu. (≥Assoc.) | -0.221 | -0.232 | -0.211 | 0.005 | -41.16 | < mp | 369.55 |
| Age | 0.113 | 0.112 | 0.113 | 4.08e-04 | 276.08 | < mp | 16554.07 |
| Age<sup>2</sup> | 0.002 | 0.002 | 0.002 | 4.53e-05 | 49.59 | < mp | 535.81 |
| Years Since Baseline | 0.010 | 0.004 | 0.015 | 0.003 | 3.59 | 3.25e-04 | 3.49 |
| PD (Case) | 0.211 | 0.178 | 0.243 | 0.017 | 12.73 | 4.21e-37 | 36.38 |
| E4 (Carrier) | 0.012 | -0.001 | 0.025 | 0.007 | 1.79 | 0.073 | 1.14 |
| PD × Age | -0.001 | -0.005 | 0.003 | 0.002 | -0.44 | 0.656 | 0.18 |
| PD × Age<sup>2</sup> | 0.001 | 4.28e-04 | 0.001 | 2.24e-04 | 3.87 | 1.07e-04 | 3.97 |
| E4 × Age | 0.005 | 0.004 | 0.007 | 0.001 | 6.75 | 1.43e-11 | 10.84 |
| E4 × Age<sup>2</sup> | 2.81e-04 | 1.06e-04 | 4.56e-04 | 8.94e-05 | 3.15 | 0.002 | 2.78 |
| PD × E4 | -0.008 | -0.077 | 0.062 | 0.036 | -0.22 | 0.826 | 0.08 |
| PD × E4 × Age | 0.020 | 0.012 | 0.028 | 0.004 | 4.75 | 2.04e-06 | 5.69 |
| PD × E4 × Age<sup>2</sup> | 0.002 | 0.001 | 0.003 | 4.79e-04 | 3.76 | 1.71e-04 | 3.77 |
| PC0 | -0.012 | -0.017 | -0.007 | 0.002 | -4.73 | 2.25e-06 | 5.65 |
| PC1 | -0.014 | -0.019 | -0.010 | 0.002 | -5.94 | 2.79e-09 | 8.55 |
| PC2 | -0.060 | -0.065 | -0.055 | 0.002 | -24.63 | 6.61e-134 | 133.18 |
| PC3 | 0.013 | 0.008 | 0.018 | 0.002 | 5.39 | 6.99e-08 | 7.16 |
| PC4 | -0.028 | -0.033 | -0.023 | 0.002 | -11.15 | 6.88e-29 | 28.16 |
| PC5 | -0.024 | -0.030 | -0.019 | 0.003 | -9.18 | 4.28e-20 | 19.37 |
| PC6 | -0.007 | -0.013 | -0.002 | 0.003 | -2.51 | 0.012 | 1.92 |
| PC7 | 1.61e-04 | -0.004 | 0.004 | 0.002 | 0.07 | 0.942 | 0.03 |
| PC8 | 0.017 | 0.013 | 0.022 | 0.002 | 7.08 | 1.42e-12 | 11.85 |
| PC9 | 0.032 | 0.028 | 0.037 | 0.002 | 14.28 | 2.75e-46 | 45.56 |
| Platform V2 | 0.305 | 0.253 | 0.358 | 0.027 | 11.47 | 1.89e-30 | 29.72 |
| Platform V3 | 0.029 | 0.010 | 0.049 | 0.010 | 2.95 | 0.003 | 2.50 |
| Platform V4 | 0.012 | 0.001 | 0.022 | 0.005 | 2.25 | 0.025 | 1.61 |
| Device: Headphones | -0.837 | -0.847 | -0.827 | 0.005 | -169.77 | < mp | 6261.07 |

Note. < mp denotes $p$ -values smaller than the machine precision (see $-\log_{10}(p)$ for more precise $p$ -values); E4 denotes APOE E4 carrier status; PD=Parkinson's disease status; PC=ancestry principal component.

**Figure 3.**
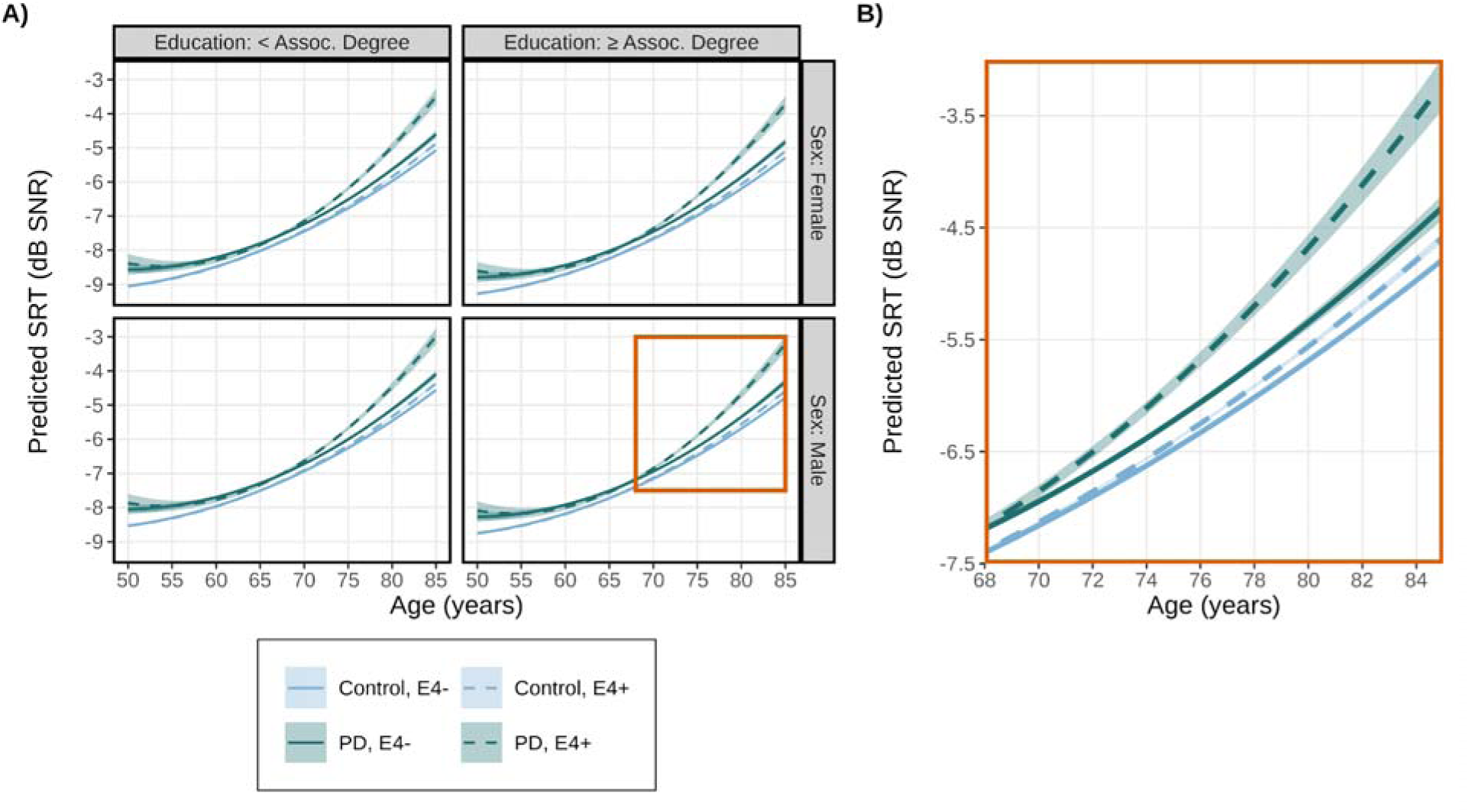
Speech-in-noise hearing loss accelerates with age, especially in *APOE* E4 carriers with Parkinson’s disease (PD). Greater speech reception thresholds (SRTs) indicate increased hearing loss. A) Conditional model predictions of SRTs were generated at fixed covariate values, assuming sample averages for ancestry principal components, the V5 genotyping platform, and laptop/computer speakers at baseline across combinations of sex and education level. B) A zoomed-in plot of predicted SRT values from males with education attainment of at least an Associates degree (emphasized covariates were arbitrarily chosen). *APOE* E4 status is designated as E4+ for carriers and E4- for non-carriers; dB=decibel; SNR=signal-to-noise ratio. Error shading is 95% CI.

## Discussion

In this large-scale study of over 240,000 23andMe Research Institute participants, we examined the independent and interactive effects of PD status and *APOE* E4 carrier status on age-related hearing loss, measured via a validated web-based SIN hearing assessment. We found that both PD and *APOE* E4 independently exacerbated age-related hearing loss, with non-linear increases in SIN hearing loss with participants’ age. Longitudinal analyses showed worsening SIN hearing loss with time independent of age. Notably, we observed a three-way interaction between PD status, *APOE* E4, and age, suggesting that the pathologies associated with PD and *APOE* E4 may interact to exacerbate hearing loss in older individuals.

### PD and Age-Related Hearing Loss

The finding that PD status interacted with age to worsen SIN performance is consistent with a growing body of evidence linking PD to auditory dysfunction (Jafari et al., 2020; Rajai Firouzabadi et al., 2026). Our results extend these findings by demonstrating that the PD-hearing loss association is not uniform across the age span but instead accelerates non-linearly with advancing age. This suggests that the pathological process linking these conditions becomes more consequential as both PD and age-related auditory decline progress. PD-related neurodegeneration may affect the auditory system at multiple levels, including the cochlea—where dopaminergic efferents modulate outer hair cell function and serve a protective role (Lendvai et al., 2011)—and the central auditory pathway, where alpha-synuclein pathology has been documented in the inferior colliculus (Seidel et al., 2015) and auditory cortex at advanced disease stages (Braak et al., 2003). Furthermore, SIN performance in older adults reflects contributions from peripheral hearing, central auditory processing, and cognitive function (Humes et al., 2012). Therefore, the PD × age^2^ interaction we observed likely reflects a combination of peripheral and central auditory decline that compounds the effects of aging, especially when PD-related pathology can contribute to both cognitive (Quadalti et al., 2023) and auditory dysfunction (Jafari et al., 2020). We propose that this peripheral and central auditory vulnerability may increase the susceptibility of PD cases to additional cognitive burden, such as those imposed by *APOE* E4.

### *APOE* E4 and Age-Related Hearing Loss

Independent of PD status, *APOE* E4 carrier status was associated with greater age-related hearing loss. This finding adds to a mixed literature on the relationship between *APOE* E4 and auditory function (Cha et al., 2024; Han et al., 2024; Jayakody et al., 2020; Tuwaig et al., 2017; Zimmermann et al., 2019); however, these studies may have been underpowered to detect the potentially small effect of *APOE* E4 on age-related hearing loss. Indeed, the linear effect of *APOE* E4 on age-related hearing loss is likely subtle as evidenced by our marginal interaction in the baseline cohort (*b*=0.002, *p*=0.055) but stronger effect in the longitudinal analysis (*b*=0.005, *p*=1.43e-11). Our study, with its large longitudinal sample, was better powered to detect *APOE* E4 interactions with age and demonstrated that the *APOE* E4 reduces SIN performance with advancing age. This result is consistent with previous work that reported a biphasic impact of the *APOE* E4 allele on age-related hearing loss (Han et al., 2024).

Given that the SIN task used here places demands on both peripheral hearing acuity and higher-order cognitive processes, the *APOE* E4 effect we observed may primarily reflect the well-documented influence of *APOE* E4 on cognitive decline rather than, or in addition to, direct effects on the peripheral auditory system (Narasimhan et al., 2024). Hearing loss is associated with increased AD neuropathology (i.e., β-amyloid and tau), especially for measures of centralized auditory processing like SIN (Chandra et al., 2025). Furthermore, a recent investigation demonstrated that hearing loss was non-uniform across different AD subtypes, with the worst hearing loss observed in a sensorimotor subtype of AD relative to a classic late-onset AD subtype with a higher polygenic risk score linked to *APOE* E4 (Lian et al., 2026). Likewise, we observed that the effect of PD on SRT performance was larger than the effect of *APOE* E4. PD-related pathology also impacts cognitive processes involved in SIN hearing ability, such as working memory and selective attention (Aarsland et al., 2021). Taken together, the loss of dopaminergic efferent projections in addition to cognitive impairment in PD may exacerbate hearing loss more than purely cognitive effects associated with *APOE* E4. Nevertheless, the effect of *APOE* E4 independently contributed to age-related hearing loss over and above PD, suggesting an independent mechanism with an underlying genetic component to hearing loss.

### The Interactive Effects of PD and *APOE* E4 on Age-Related SIN Hearing Loss

In participants with at least two longitudinal SIN hearing assessments, we observed a three-way interaction between PD status, *APOE* E4, and age^2^. This result suggests that the neuropathologies resulting from PD and *APOE* E4 carrier status may interact to exacerbate age-related SIN hearing loss. Because the loss of dopaminergic neurons affects both peripheral and centralized hearing pathways (Jafari et al., 2020; Rajai Firouzabadi et al., 2026), as well as cognitive functions critical to SIN hearing ability (Aarsland et al., 2021), individuals with PD are particularly vulnerable to additional age-related hearing loss. In contrast, *APOE* E4 associated pathology primarily acts on centralized hearing processes (Chandra et al., 2025; Tuwaig et al., 2017), although the influence of *APOE* E4 on peripheral auditory pathways cannot be ruled out. *APOE* E4 carriers with PD report greater cognitive and memory impairment (Kmiecik et al., 2025), and concomitant pathologies, such alpha-synuclein and amyloid/tau burden, show accelerated cognitive decline (Quadalti et al., 2023; Szwedo et al., 2022). Therefore, the combination of both PD and *APOE* E4 likely affects cognition and auditory pathways at multiple levels, resulting in exacerbated SIN hearing loss for *APOE* E4 carriers with PD.

The three-way interaction between PD, *APOE* E4, and age^2^ was observed in the longitudinal cohort and not the larger baseline cohort. These differing results likely reflect the advantages of repeated-measures to effectively partition stable between-subject variance via random effects. Additionally, the participants in the longitudinal cohort are highly-motivated participants by virtue of their repeated participation, sometimes across several years, in the SIN hearing assessment and related surveys. Therefore, we hypothesize that the longitudinal subsample was likely more homogeneous and motivated to perform well relative to the larger cross-sectional baseline sample, thus increasing our outcome’s SNR to detect an effect.

## Clinical Implications

Our findings reinforce the growing recognition that hearing loss is a clinically meaningful non-motor feature of PD and identify *APOE* E4 carriers as an additional group at elevated risk for accelerated age-related hearing decline. Individuals with PD and particularly those who also carry the *APOE* E4 allele may represent a high-priority population for hearing loss screening and intervention. The non-linear acceleration of hearing loss with age in these groups implies that earlier identification and management of hearing difficulties could mitigate downstream effects on communication, social engagement, and cognitive function. In one of the largest studies examining SIN hearing and incident dementia using a digit triplets test, Stevenson et al. (2022) found that depressive symptoms and social isolation combined accounted for less than 7% of the excess dementia risk associated with SIN hearing impairment. This suggests that the association between SIN hearing impairment and dementia operates largely through pathways independent of mood and social engagement, and that hearing loss itself represents a direct and actionable target for intervention. However, more work is needed to disentangle the causal relationship underlying hearing loss and downstream effects like dementia, for which PD and *APOE* E4 are pertinent risk factors.

The web-based SIN assessment used in this study offers a scalable approach to hearing screening that could be integrated into routine PD care, potentially identifying hearing decline before it is self-reported. Although randomized controlled trials of hearing interventions and PD risk are currently absent, the convergence of evidence from the ACHIEVE trial (Pike et al., 2025), observational studies (Jafari et al., 2020; Neilson et al., 2024; Rajai Firouzabadi et al., 2026), and the present findings provides a strong rationale for such trials. Given that PD and *APOE* E4 appear to affect hearing through partially distinct, though interacting, mechanisms, hearing interventions may provide benefits that complement disease-modifying therapies targeting either PD or AD pathology specifically. Given the complex interplay between peripheral and central auditory processing, and the effects of neurodegeneration on these pathways, hearing aid interventions alone may not sufficiently restore hearing loss from auditory signal amplification (Johnson et al., 2021). Therefore, additional interventions should be also considered, including “smart” hearing aids and auditory-based cognitive-behavioral therapies (Levett et al., 2025).

## Limitations

Several limitations should be considered when interpreting these results. PD diagnosis was determined by self-report. Although self-reported PD status at 23andMe has excellent concordance with clinical exams (Dorsey et al., 2015; Myers et al., 2022; Winslow et al., 2018) and we excluded participants who revised their PD diagnosis, misclassification of PD status was possible. Pure-tone audiometry was not performed, precluding adjustment for peripheral hearing loss and limiting our ability to disentangle peripheral hearing loss from central contributions to SIN deficits (Humes et al., 2012). The 23andMe Research Institute cohort is predominantly of European ancestry and represents a self-selected population enriched for higher educational attainment, which may limit generalizability. We excluded *LRRK2* p.G2019S and *GBA1* p.N409S carriers to focus on idiopathic PD, and our findings may not generalize to these genetic subtypes, which have distinct clinical profiles and may relate to hearing loss differently (Kmiecik et al., 2024, 2025; Sosero & Gan-Or, 2023). Given that our sample comprised only 6–7% of *APOE* E4 homozygous carriers, we did not distinguish between *APOE* E4 haplotypes. Given the increased risk of Alzheimer’s disease pathology in *APOE* E4 homozygous carriers (Fortea et al., 2024), future, adequately powered studies would be well served to examine the impact of *APOE* E4 haplotypes on age-related hearing loss. Finally, the detection of statistical interactions is scale-dependent (González & Cox, 2007); our finding is conditional on the decibel scale, the conventional metric for SIN performance.

## Conclusion

In the largest study to date examining the joint effects of PD and *APOE* E4 on SIN hearing loss, we demonstrate that both PD and *APOE* E4 interact to exacerbate age-related decline in SIN performance, with effects that accelerate non-linearly with age. These findings support the inclusion of hearing screening in routine PD care and highlight individuals with PD and *APOE* E4 carriers as priority populations for hearing intervention, with the potential to mitigate compounding effects on communication and cognition in aging. Future work would be well served to examine hearing interventions in PD, such as the dispensation of hearing aids and auditory therapies, and their effects on quality of life, cognition, and progression of motor and non-motor symptoms in PD.

## Data Availability

There are restrictions to the availability of the 23andMe data due to 23andMe consent and privacy guidelines. These data will not be made available. No custom code or software was generated as part of the study. Details of all software packages used for data processing and analysis may be found in the Methods.

## Acknowledgements

We would like to thank the research participants and employees of 23andMe Research Institute for making this work possible and The Michael J. Fox Foundation for Parkinson’s Research for funding this research. The following members of the 23andMe Research Team contributed to this study:

Noura S. Abul-Husn, Aditya Ambati, James R. Ashenhurst, Robert K. Bell, Rebecca M. K. Berns, Kahsaia de Brito, Stacey B. Detweiler, Emily DelloRusso, Sayantan Das, William A. Freyman, Chris German, Éadaoin Harney, Alejandro Hernandez-Chavez, Barry Hicks, David A. Hinds, Ethan M. Jewett, Bertram L. Koelsch, Alan Kwong, Katelyn Kukar Bond, Alisa P. Lehman, Keng-Han Lin, Steven J. Micheletti, Alexander Moran, G. David Poznik, Emily M. Rios, Shubham Saini, Anjali J. Shastri, Qiaojuan Jane Su, Ruth I. Tennen, Vinh Tran, Joyce Y. Tung, Peter R. Wilton, R. Ryanne Wu, Jingran Wen

## Ethics Statement

Participants provided informed consent and volunteered to participate in the research online, under a protocol approved by the external AAHRPP-accredited Salus IRB (https://www.versiticlinicaltrials.org/salusirb). Accordingly, only participants from the United States and United Kingdom were included in analyses given legal/regulatory compliance policies.

## References

1. 23andMe, Inc. (2022). Ancestry Composition: 23andMe’s State-of-the-Art Geographic Ancestry Analysis. https://www.23andme.com/ancestry-composition-guide/

2. 23andMe Research Team. (2024). Overview of 23andMe GWAS release: R8_g1 [White Paper]. 23andMe, Inc. https://permalinks.23andme.com/pdf/23andMe-technical-overview-gwas-r8_g1.pdf

3. Aarsland, D., Batzu, L., Halliday, G. M., Geurtsen, G. J., Ballard, C., Ray Chaudhuri, K., & Weintraub, D. (2021). Parkinson disease-associated cognitive impairment. Nature Reviews Disease Primers, 7(1), 47. 10.1038/s41572-021-00280-3

4. Arel-Bundock, V., Greifer, N., & Heiss, A. (2024). How to Interpret Statistical Models Using marginaleffects for R and Python. Journal of Statistical Software, 111(9), 1–32. 10.18637/jss.v111.i09

5. Billings, C. J., Olsen, T. M., Charney, L., Madsen, B. M., & Holmes, C. E. (2023). Speech-in-Noise Testing: An Introduction for Audiologists. Seminars in Hearing, 45(1), 55–82. 10.1055/s-0043-1770155

6. Braak, H., Tredici, K. D., Rüb, U., de Vos, R. A. I., Jansen Steur, E. N. H., & Braak, E. (2003). Staging of brain pathology related to sporadic Parkinson’s disease. Neurobiology of Aging, 24(2), 197–211. 10.1016/S0197-4580(02)00065-9

7. Cha, C.-H., Lin, T.-K., Wu, C.-N., Yang, C.-H., Huang, Y.-W., & Hwang, C.-F. (2024). Relationship of Hearing Loss to Parkinson’s Disease, Dementia, and APOE Genotype in Adults. Medicina, 60(5). 10.3390/medicina60050703

8. Chandra, A., Levett, B. A., Waters, S., Proitsi, P., Liu, Y., Hardy, C. J. D., Warren, J. D., & Marshall, C. R. (2025). Evaluating the link between hearing loss and Alzheimer’s disease neuropathology: A systematic review and meta-analysis. Neurobiology of Aging, 154, 92–102. 10.1016/j.neurobiolaging.2025.07.003

9. De Sousa, K. C., Swanepoel, D. W., Moore, D. R., & Smits, C. (2018). A Smartphone National Hearing Test: Performance and Characteristics of Users. American Journal of Audiology, 27(3S), 448–454. 10.1044/2018_AJA-IMIA3-18-0016

10. Dorsey, E. R., Darwin, K. C., Mohammed, S., Donohue, S., Tethal, A., Achey, M. A., Ward, S., Caughey, E., Conley, E. D., Eriksson, N., & Ravina, B. (2015). Virtual research visits and direct-to-consumer genetic testing in Parkinson’s disease. DIGITAL HEALTH, 1, 2055207615592998. 10.1177/2055207615592998

11. Durand, E. Y., Do, C. B., Mountain, J. L., & Macpherson, J. M. (2014). Ancestry Composition: A Novel, Efficient Pipeline for Ancestry Deconvolution (p. 010512). bioRxiv. 10.1101/010512

12. Fortea, J., Pegueroles, J., Alcolea, D., Belbin, O., Dols-Icardo, O., Vaqué-Alcázar, L., Videla, L., Gispert, J. D., Suárez-Calvet, M., Johnson, S. C., Sperling, R., Bejanin, A., Lleó, A., & Montal, V. (2024). APOE4 homozygozity represents a distinct genetic form of Alzheimer’s disease. Nature Medicine. 10.1038/s41591-024-02931-w

13. González, A. B. de, & Cox, D. R. (2007). Interpretation of interaction: A review. The Annals of Applied Statistics, 1(2), 371–385. 10.1214/07-AOAS124

14. Han, J. S., Yoo, S. goo, Lee, S. jung, Lee, H. J., Choi, I. Y., & Park, K. H. (2024). The biphasic impact of apolipoprotein E ε4 allele on age-related hearing loss. Scientific Reports, 14(1), 21420. 10.1038/s41598-024-71774-9

15. Harris, C. R., Millman, K. J., van der Walt, S. J., Gommers, R., Virtanen, P., Cournapeau, D., Wieser, E., Taylor, J., Berg, S., Smith, N. J., Kern, R., Picus, M., Hoyer, S., van Kerkwijk, M. H., Brett, M., Haldane, A., del Río, J. F., Wiebe, M., Peterson, P., … Oliphant, T. E. (2020). Array programming with NumPy. Nature, 585(7825), 357–362. 10.1038/s41586-020-2649-2

16. Humes, L. E., Dubno, J. R., Gordon-Salant, S., Lister, J. J., Cacace, A. T., Cruickshanks, K. J., Gates, G. A., Wilson, R. H., & Wingfield, A. (2012). Central Presbycusis: A Review and Evaluation of the Evidence. Journal of the American Academy of Audiology, 23(8), 635–666. 10.3766/jaaa.23.8.5

17. Jafari, Z., Kolb, B. E., & Mohajerani, M. H. (2020). Auditory Dysfunction in Parkinson’s Disease. Movement Disorders, 35(4), 537–550. 10.1002/mds.28000

18. Jayakody, D. M. P., Menegola, H. K., Yiannos, J. M., Goodman-Simpson, J., Friedland, P. L., Taddei, K., Laws, S. M., Weinborn, M., Martins, R. N., & Sohrabi, H. R. (2020). The Peripheral Hearing and Central Auditory Processing Skills of Individuals With Subjective Memory Complaints. Frontiers in Neuroscience, 14. 10.3389/fnins.2020.00888

19. Johnson, J. C. S., Marshall, C. R., Weil, R. S., Bamiou, D.-E., Hardy, C. J. D., & Warren, J. D. (2021). Hearing and dementia: From ears to brain. Brain, 144(2), 391–401. 10.1093/brain/awaa429

20. Kloske, C. M., Belloy, M. E., Blue, E. E., Bowman, G. R., Carrillo, M. C., Chen, X., Chiba-Falek, O., Davis, A. A., Paolo, G. D., Garretti, F., Gate, D., Golden, L. R., Heinecke, J. W., Herz, J., Huang, Y., Iadecola, C., Johnson, L. A., Kanekiyo, T., Karch, C. M., … Holtzman, D. M. (2024). Advancements in APOE and dementia research: Highlights from the 2023 AAIC Advancements: APOE conference. Alzheimer’s & Dementia, 20(9), 6590–6605. 10.1002/alz.13877

21. Kmiecik, M. J., Holmes, M. V., Fontanillas, P., Riboldi, G. M., Schneider, R. B., Shi, J., Guan, A., Tat, S., Micheletti, S., Stagaman, K., Gottesman, J., Hinds, D. A., Tung, J. Y., Team, 23andMe Research, Aslibekyan, S., & Norcliffe-Kaufmann, L. (2025). Genetic Modifiers of Parkinson’s Disease: A Case–Control Study. Annals of Clinical and Translational Neurology, 12(12), 2482–2494. 10.1002/acn3.70176

22. Kmiecik, M. J., Micheletti, S., Coker, D., Heilbron, K., Shi, J., Stagaman, K., Filshtein Sonmez, T., Fontanillas, P., Shringarpure, S., Wetzel, M., Rowbotham, H. M., Cannon, P., Shelton, J. F., Hinds, D. A., Tung, J. Y., 23andMe Research Team, Holmes, M. V., Aslibekyan, S., & Norcliffe-Kaufmann, L. (2024). Genetic analysis and natural history of Parkinson’s disease due to the LRRK2 G2019S variant. Brain, 147(6), 1996–2008. 10.1093/brain/awae073

23. Lai, S.-W., Liao, K.-F., Lin, C.-L., Lin, C.-C., & Sung, F.-C. (2014). Hearing loss may be a non-motor feature of Parkinson’s disease in older people in Taiwan. European Journal of Neurology, 21(5), 752–757. 10.1111/ene.12378

24. Lendvai, B., Halmos, G. B., Polony, G., Kapocsi, J., Horváth, T., Aller, M., Sylvester Vizi, E., & Zelles, T. (2011). Chemical neuroprotection in the cochlea: The modulation of dopamine release from lateral olivocochlear efferents. Neurochemistry International, 59(2), 150–158. 10.1016/j.neuint.2011.05.015

25. Levett, B. A., Chandra, A., Jiang, J., Koohi, N., Sharrad, D., Core, L. B., Johnson, J. C. S., Tutton, M., Green, T., Jayakody, D. M. P., Yu, J.-T., Leroi, I., Marshall, C. R., Bamiou, D.-E., Hardy, C. J. D., & Warren, J. D. (2025). Hearing impairment and dementia: Cause, catalyst or consequence? Journal of Neurology, 272(6), 402. 10.1007/s00415-025-13140-x

26. Lian, J., Fan, Z., Petrazzini, B. O., Fan, W., Rao, S., Yang, Q., Zeng, G., Ahmed, N., Tabassi Mofrad, F., Wamil, M., & Rahimi, K. (2026). Subtyping Alzheimer’s disease and Parkinson’s disease using longitudinal electronic health records. Nature Aging, 1–14. 10.1038/s43587-026-01085-3

27. Lin, F. R. (2024). Age-Related Hearing Loss. New England Journal of Medicine, 390(16), 1505–1512. 10.1056/NEJMcp2306778

28. Lin, F. R., Pike, J. R., Albert, M. S., Arnold, M., Burgard, S., Chisolm, T., Couper, D., Deal, J. A., Goman, A. M., Glynn, N. W., Gmelin, T., Gravens-Mueller, L., Hayden, K. M., Huang, A. R., Knopman, D., Mitchell, C. M., Mosley, T., Pankow, J. S., Reed, N. S., … Coresh, J. (2023). Hearing intervention versus health education control to reduce cognitive decline in older adults with hearing loss in the USA (ACHIEVE): A multicentre, randomised controlled trial. The Lancet, 402(10404), 786–797. 10.1016/S0140-6736(23)01406-X

29. Livingston, G., Huntley, J., Sommerlad, A., Ames, D., Ballard, C., Banerjee, S., Brayne, C., Burns, A., Cohen-Mansfield, J., Cooper, C., Costafreda, S. G., Dias, A., Fox, N., Gitlin, L. N., Howard, R., Kales, H. C., Kivimäki, M., Larson, E. B., Ogunniyi, A., … Mukadam, N. (2020). Dementia prevention, intervention, and care: 2020 report of the Lancet Commission. The Lancet, 396(10248), 413–446. 10.1016/S0140-6736(20)30367-6

30. Myers, T. L., Augustine, E. F., Baloga, E., Daeschler, M., Cannon, P., Rowbotham, H., Chanoff, E., Team, 23andMe Research, Jensen-Roberts, S., Soto, J., Holloway, R. G., Marras, C., Tanner, C. M., Dorsey, E. R., & Schneider, R. B. (2022). Recruitment for Remote Decentralized Studies in Parkinson’s Disease. Journal of Parkinson’s Disease, 12(1), 371–380. 10.3233/JPD-212935

31. Narasimhan, S., Holtzman, D. M., Apostolova, L. G., Cruchaga, C., Masters, C. L., Hardy, J., Villemagne, V. L., Bell, J., Cho, M., & Hampel, H. (2024). Apolipoprotein E in Alzheimer’s disease trajectories and the next-generation clinical care pathway. Nature Neuroscience, 27(7), 1236–1252. 10.1038/s41593-024-01669-5

32. Neilson, L. E., Reavis, K. M., Wiedrick, J., & Scott, G. D. (2024). Hearing Loss, Incident Parkinson Disease, and Treatment With Hearing Aids. JAMA Neurology, 81(12), 1295. 10.1001/jamaneurol.2024.3568

33. Ning, P., Mu, X., Guo, X., & Li, R. (2024). Hearing loss is not associated with risk of Parkinson’s disease: A Mendelian randomization study. Heliyon, 10(11). 10.1016/j.heliyon.2024.e32533

34. Patil, I., Makowski, D., Ben-Shachar, M. S., Wiernik, B. M., Bacher, E., & Lüdecke, D. (2022). datawizard: An R Package for Easy Data Preparation and Statistical Transformations. Journal of Open Source Software, 7(78), 4684. 10.21105/joss.04684

35. Pike, J. R., Huang, A. R., Reed, N. S., Arnold, M., Chisolm, T., Couper, D., Deal, J. A., Glynn, N. W., Goman, A. M., Hayden, K. M., Mitchell, C. M., Pankow, J. S., Sanchez, V., Sullivan, K. J., Tan, N. S., Coresh, J., & Lin, F. R. (2025). Cognitive benefits of hearing intervention vary by risk of cognitive decline: A secondary analysis of the ACHIEVE trial. Alzheimer’s & Dementia, 21(5), e70156. 10.1002/alz.70156

36. Potgieter, J.-M., Swanepoel, D. W., Myburgh, H. C., Hopper, T. C., & Smits, C. (2016). Development and validation of a smartphone-based digits-in-noise hearing test in South African English. International Journal of Audiology, 55(7), 405–411. 10.3109/14992027.2016.1172269

37. Python Software Foundation. (2016). Python (Version Version 3.12.5) [Computer software]. https://www.python.org/

38. Quadalti, C., Palmqvist, S., Hall, S., Rossi, M., Mammana, A., Janelidze, S., Dellavalle, S., Mattsson-Carlgren, N., Baiardi, S., Stomrud, E., Hansson, O., & Parchi, P. (2023). Clinical effects of Lewy body pathology in cognitively impaired individuals. Nature Medicine, 29(8), 1964–1970. 10.1038/s41591-023-02449-7

39. R Core Team. (2024). R: A language and environment for statistical computing. [Computer software]. R Foundation for Statistical Computing. Vienna, Austria. https://www.R-project.org/

40. Rajai Firouzabadi, S., Mohammadi, I., Golsorkh, H., Mahdavi, S., Sadeghzadeh, S., Rezaei, K., & Salari, M. (2026). Hearing Loss in Parkinson’s Disease: A Systematic Review and Meta-Analysis. Movement Disorders, n/a(n/a). 10.1002/mds.70206

41. Readman, M. R., Wang, Y., Wan, F., Fairman, I., Linkenauger, S. A., Crawford, T. J., & Plack, C. J. (2025). Speech-in-noise hearing impairment is associated with increased risk of Parkinson’s: A UK biobank analysis. Parkinsonism & Related Disorders, 131, 107219. 10.1016/j.parkreldis.2024.107219

42. Rigby, R. A., & Stasinopoulos, D. M. (2005). Generalized Additive Models for Location, Scale and Shape. Journal of the Royal Statistical Society Series C: Applied Statistics, 54(3), 507–554. 10.1111/j.1467-9876.2005.00510.x

43. Rosal, A. E., Martin, S. L., & Strafella, A. P. (2025). The role of Apolipoprotein E4 on cognitive impairment in Parkinson’s disease and Parkinsonisms. Frontiers in Neuroscience, 19. 10.3389/fnins.2025.1515374

44. Seidel, K., Mahlke, J., Siswanto, S., Krüger, R., Heinsen, H., Auburger, G., Bouzrou, M., Grinberg, L. T., Wicht, H., Korf, H.-W., den Dunnen, W., & Rüb, U. (2015). The Brainstem Pathologies of Parkinson’s Disease and Dementia with Lewy Bodies. Brain Pathology, 25(2), 121–135. 10.1111/bpa.12168

45. Simonet, C., Bestwick, J., Jitlal, M., Waters, S., Ben-Joseph, A., Marshall, C. R., Dobson, R., Marrium, S., Robson, J., Jacobs, B. M., Belete, D., Lees, A. J., Giovannoni, G., Cuzick, J., Schrag, A., & Noyce, A. J. (2022). Assessment of Risk Factors and Early Presentations of Parkinson Disease in Primary Care in a Diverse UK Population. JAMA Neurology, 79(4), 359–369. 10.1001/jamaneurol.2022.0003

46. Sosero, Y. L., & Gan-Or, Z. (2023). LRRK2 and Parkinson’s disease: From genetics to targeted therapy. Annals of Clinical and Translational Neurology, 10(6), 850–864. 10.1002/acn3.51776

47. Stasinopoulos, D. M., & Rigby, R. A. (2008). Generalized Additive Models for Location Scale and Shape (GAMLSS) in R. Journal of Statistical Software, 23, 1–46. 10.18637/jss.v023.i07

48. Stevenson, J. S., Clifton, L., Kuźma, E., & Littlejohns, T. J. (2022). Speech-in-noise hearing impairment is associated with an increased risk of incident dementia in 82,039 UK Biobank participants. Alzheimer’s & Dementia, 18(3), 445–456. 10.1002/alz.12416

49. Szwedo, A. A., Dalen, I., Pedersen, K. F., Camacho, M., Bäckström, D., Forsgren, L., Tzoulis, C., Winder-Rhodes, S., Hudson, G., Liu, G., Scherzer, C. R., Lawson, R. A., Yarnall, A. J., Williams-Gray, C. H., Macleod, A. D., Counsell, C. E., Tysnes, O.-B., Alves, G., Maple-Grødem, J., & Collaboration, P. I. C. (2022). GBA and APOE Impact Cognitive Decline in Parkinson’s Disease: A 10-Year Population-Based Study. Movement Disorders, 37(5), 1016–1027. 10.1002/mds.28932

50. The pandas development, team. (2026). pandas-dev/pandas: Pandas [Computer software]. Zenodo. 10.5281/zenodo.18328522

51. Tuwaig, M., Savard, M., Jutras, B., Poirier, J., Collins, D. L., Rosa-Neto, P., Fontaine, D., & Breitner, J. C. S. (2017). Deficit in Central Auditory Processing as a Biomarker of Pre-Clinical Alzheimer’s Disease. Journal of Alzheimer’s Disease, 60(4), 1589–1600. 10.3233/JAD-170545

52. Wickham, H., Averick, M., Bryan, J., Chang, W., McGowan, L. D., François, R., Grolemund, G., Hayes, A., Henry, L., Hester, J., Kuhn, M., Pedersen, T. L., Miller, E., Bache, S. M., Müller, K., Ooms, J., Robinson, D., Seidel, D. P., Spinu, V., … Yutani, H. (2019). Welcome to the Tidyverse. Journal of Open Source Software, 4(43), 1686. 10.21105/joss.01686

53. Winslow, A. R., Hyde, C. L., Wilk, J. B., Eriksson, N., Cannon, P., Miller, M. R., & Hirst, W. D. (2018). Self-report data as a tool for subtype identification in genetically-defined Parkinson’s Disease. Scientific Reports, 8(1), Article 1. 10.1038/s41598-018-30843-6

54. Yu, R.-C., Proctor, D., Soni, J., Pikett, L., Livingston, G., Lewis, G., Schilder, A., Bamiou, D., Mandavia, R., Omar, R., Pavlou, M., Lin, F., Goman, A. M., & Gonzalez, S. C. (2024). Adult-onset hearing loss and incident cognitive impairment and dementia – A systematic review and meta-analysis of cohort studies. Ageing Research Reviews, 98, 102346. 10.1016/j.arr.2024.102346

55. Zhang, H., Chen, K., Gao, T., Yan, Y., Liu, Y., Liu, Y., Zhu, K., Qi, J., Zheng, C., Wang, T., & Zeng, P. (2025). Establishing a robust triangulation framework to explore the relationship between hearing loss and Parkinson’s disease. Npj Parkinson’s Disease, 11(1), 5. 10.1038/s41531-024-00861-5

56. Zimmermann, J., Alain, C., & Butler, C. (2019). Impaired memory-guided attention in asymptomatic APOE4 carriers. Scientific Reports, 9(1), 8138. 10.1038/s41598-019-44471-1

