## Supplementary Materials for "Age-Related Speech-in-Noise Hearing Loss in Parkinson’s Disease and *APOE* E4 Carriers"

Last modified: 2026-05-27 16:42:38

### Table of contents

**1 Genotyping** **1**

**2 Principal Components of Ancestry** **2**

**3 References** **7**

### 1 Genotyping

DNA extraction and genotyping were performed on saliva samples by Clinical Laboratory Improvement Amendments-certified and College of American Pathologists-accredited clinical laboratories of Laboratory Corporation of America. Samples were genotyped on one of five genotyping platforms. The V1 and V2 platforms were variants of the Illumina HumanHap550+ BeadChip and contained a total of approximately 560,000 single nucleotide polymorphisms (SNPs), including about 25,000 custom SNPs selected by 23andMe. The V3 platform was based on the Illumina OmniExpress + BeadChip and contained a total of about 950,000 SNPs and custom content to improve the overlap

Table S1: Model comparison statistics for baseline cross-sectional models.

| Model | Family | N | df | Log-Likelihood | AIC | BIC | ΔAIC | ΔBIC |
|---|---|---|---|---|---|---|---|---|
| GAMLSS | Skew-t (ST1) | 243981 | 31 | -529228.3 | 1058518 | 1058841 | 0.0 | 0.0 |
| GAMLSS | Skew-Normal (SN1) | 243981 | 30 | -580881.2 | 1161822 | 1162135 | 103304.0 | 103293.5 |
| OLS | Linear (Gaussian) | 243981 | 28 | -612204.5 | 1224467 | 1224769 | 165948.4 | 165927.6 |

*Note.* GAMLSS (Generalized Additive Models for Location, Scale and Shape) models with skew-normal and skew-t distributional families compared against ordinary least squares (OLS) linear regression.

with our V2 array. The V4 platform is a fully custom array and includes a lower redundancy subset of V2 and V3 SNPs with additional coverage of lower-frequency coding variation, and about 570,000 SNPs. The V5 platform is an Illumina Infinium Global Screening Array of about 640,000 SNPs supplemented with about 50,000 SNPs of custom content.

Greater than 99% of the genotyping results agreed with the Sanger sequencing results for *LRRK2* p.G2019S, *GBA1* p.N409S, and the two SNPs (rs429358 and rs7412) used to determine the number of *APOE* E4 alleles. Likewise, these variants exhibited >99% reproducibility and repeatability on the genotyping platform (23andMe, 2023).

### 2 Principal Components of Ancestry

Principal components (PCs) were derived from a principal component analysis performed on one million randomly selected participants from 23andMe's Research Cohort using 63,528 high quality genotyped variants present across all five genotyping platforms. Loadings for participants not included in the analysis were obtained by projection, combining the eigenvectors of the analysis and the SNP weights. Genetic ancestry PCs were then normalized ($M$=0, $SD$=1) separately for each model.

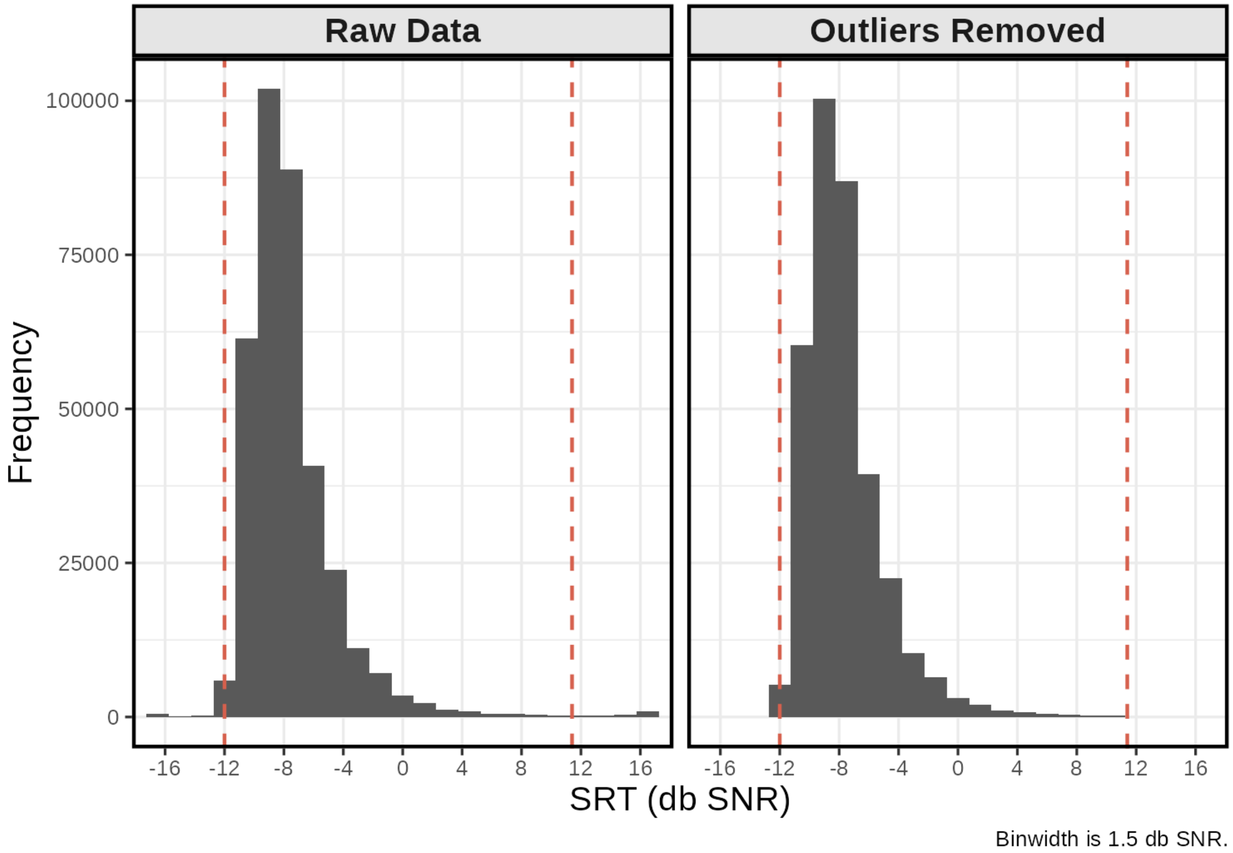

Figure S1: Distribution of speech reception thresholds (SRT) measured in decibels (dB) of signal-to-noise ratio (SNR). Participants with SRT values outside the 99th percentile (i.e., in the bottom 0.5% and top 0.5% of the SRT distribution) were identified as outliers and excluded. Left: SRT distribution prior to 99% thresholding. Right: SRT distribution following 99% thresholding outlier exclusion. Distributions include longitudinal isntances.

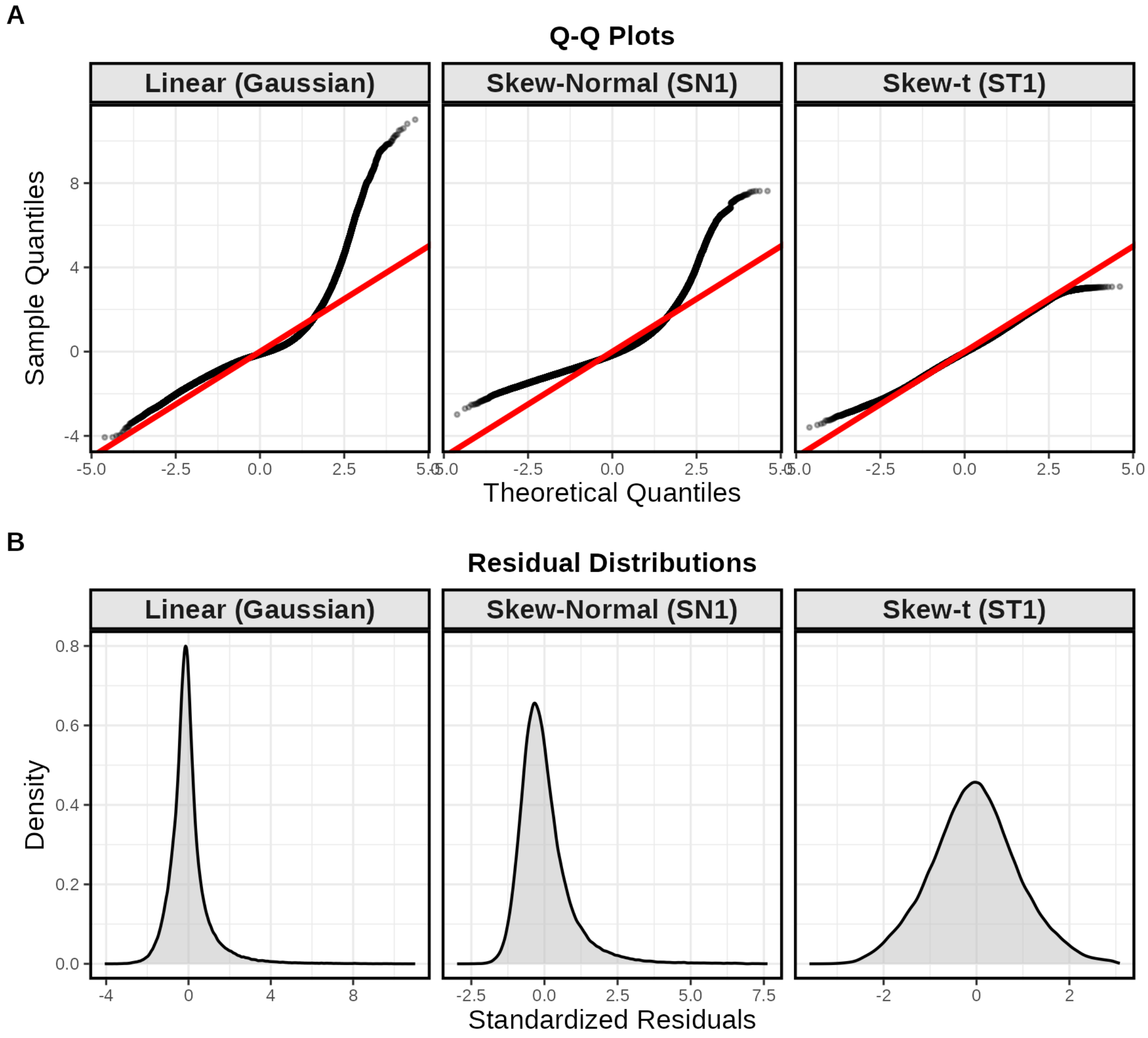

Figure S2: Model diagnostic plots of baseline cross-sectional models. A) Normal Quantile-Quantile (Q-Q) plots of residuals for linear regression, and GAMLSS models with skew-normal and skew-t distributional families. B) Density plots of of standardized residulas for the three models listed in A.

Table S2: Cross-sectional GAMLSS: Unweighted Participant Characteristics at Baseline.

| | | Controls | | PD Cases | |
|---|---|---|---|---|---|
| Variable | Category | E4- | E4+ | E4- | E4+ |
| N | | 178,402 | 61,218 | 3,346 | 1,015 |
| Age | | 64.72 (0.020) | 64.26 (0.033) | 69.51 (0.126) | 68.58 (0.224) |
| Sex | Male | 43.1% (0.1%) | 42.7% (0.2%) | 59.1% (0.8%) | 57.9% (1.5%) |
| Education | Assoc. degree | 69.7% (0.1%) | 70.2% (0.2%) | 75.7% (0.7%) | 78.8% (1.3%) |
| Genetic Ancestry | European | 85.7% (0.1%) | 87.8% (0.1%) | 91.4% (0.5%) | 92.5% (0.8%) |
| Age at PD diagnosis | | - | - | 63.52 (0.172) | 63.23 (0.295) |
| APOE E4 status | Homozygous | - | 7.3% (0.1%) | - | 7.4% (0.8%) |
| Speech reception threshold (dB) | | -7.76 (0.006) | -7.80 (0.011) | -6.78 (0.051) | -6.78 (0.099) |
| Device used for test | Headphones | 24.7% (0.1%) | 25.6% (0.2%) | 22.7% (0.7%) | 26.9% (1.4%) |
| Background noise level (0-5) | | 0.99 (0.003) | 0.99 (0.005) | 1.01 (0.020) | 0.99 (0.037) |
| Difficulty hearing in noise | A lot of difficulty | 13.5% (0.1%) | 13.5% (0.1%) | 18.0% (0.7%) | 17.6% (1.2%) |
| Self-reported hearing problems | Yes | 36.7% (0.1%) | 36.5% (0.2%) | 44.0% (0.9%) | 43.1% (1.6%) |
| Degree of hearing loss | None/Mild | 58.7% (0.2%) | 59.4% (0.3%) | 52.5% (1.3%) | 55.4% (2.5%) |
| Diagnosed with hearing loss | Yes | 92.0% (0.1%) | 91.2% (0.2%) | 94.2% (0.7%) | 93.2% (1.4%) |
| Age at hearing loss diagnosis | | 52.90 (0.084) | 52.07 (0.145) | 55.02 (0.515) | 54.77 (0.976) |
| Use cochlear implant | Yes | 0.1% (0.0%) | 0.1% (0.0%) | 0.2% (0.1%) | Masked |
| Age started cochlear implant | | 57.91 (0.912) | 57.55 (1.508) | 67.86 (3.003) | Masked |
| Use hearing aid | Yes | 11.2% (0.1%) | 10.7% (0.1%) | 18.1% (0.7%) | 17.4% (1.2%) |
| Age started hearing aid | | 62.49 (0.081) | 61.77 (0.142) | 64.07 (0.432) | 63.36 (0.865) |
| Use assistive listening device | Yes | 0.5% (0.0%) | 0.5% (0.0%) | 0.7% (0.1%) | 1.0% (0.3%) |
| Age started assistive device | | 60.61 (0.409) | 60.16 (0.768) | 64.50 (1.776) | 59.64 (2.165) |
| No hearing devices | Yes | 88.4% (0.1%) | 88.9% (0.1%) | 81.4% (0.7%) | 81.9% (1.2%) |
| Ménière’s disease | Yes | 2.0% (0.0%) | 2.0% (0.1%) | 2.1% (0.3%) | 2.7% (0.5%) |
| Vertigo | Yes | 37.1% (0.1%) | 37.1% (0.2%) | 41.2% (0.9%) | 40.2% (1.6%) |
| Tinnitus | Yes | 33.6% (0.1%) | 33.9% (0.2%) | 37.5% (0.9%) | 34.3% (1.5%) |
| Tinnitus severity (0-10) | | 2.78 (0.011) | 2.82 (0.019) | 2.70 (0.072) | 2.65 (0.138) |

*Note.* Categorical measures presented as % (SE), continuous as Mean (SEM). E4+ indicates APOE E4 carriers, E4- non-carriers. Indented items are contingent follow-ups. Statistics with n<5 masked for privacy; dashes indicate not applicable; PD=Parkinson’s disease; SRT=speech reception threshold..

Table S3: Longitudinal GAMLSS: Unweighted Participant Characteristics at Baseline.

| | | Controls | | PD Cases | |
|---|---|---|---|---|---|
| Variable | Category | E4- | E4+ | E4- | E4+ |
| N | | 26,990 | 9,252 | 1,131 | 303 |
| Age | | 67.27 (0.049) | 66.72 (0.082) | 70.16 (0.198) | 69.43 (0.387) |
| Sex | Male | 45.8% (0.3%) | 44.8% (0.5%) | 59.2% (1.5%) | 58.4% (2.8%) |
| Education | Assoc. degree | 73.9% (0.3%) | 74.6% (0.5%) | 77.3% (1.2%) | 83.8% (2.1%) |
| Genetic Ancestry | European | 88.4% (0.2%) | 90.7% (0.3%) | 91.1% (0.8%) | 94.7% (1.3%) |
| Age at PD diagnosis | | - | - | 64.77 (0.269) | 64.65 (0.508) |
| APOE E4 status | Homozygous | - | 7.4% (0.3%) | - | 5.9% (1.4%) |
| Speech reception threshold (dB) | | -7.56 (0.016) | -7.61 (0.028) | -6.80 (0.084) | -6.81 (0.176) |
| Device used for test | Speaker | 74.6% (0.3%) | 73.5% (0.5%) | 77.1% (1.2%) | 71.3% (2.6%) |
| Background noise level (0-5) | | 0.91 (0.007) | 0.92 (0.011) | 0.99 (0.033) | 0.94 (0.069) |
| Difficulty hearing in noise | A lot of difficulty | 15.5% (0.2%) | 15.1% (0.4%) | 17.8% (1.1%) | 18.3% (2.2%) |
| Self-reported hearing problems | Yes | 41.4% (0.3%) | 41.3% (0.5%) | 44.6% (1.5%) | 44.0% (3.0%) |
| Degree of hearing loss | None/Mild | 60.9% (0.4%) | 62.4% (0.8%) | 56.5% (2.1%) | 59.9% (4.1%) |
| Diagnosed with hearing loss | Yes | 91.0% (0.3%) | 89.3% (0.5%) | 92.1% (1.3%) | 89.5% (2.8%) |
| Age at hearing loss diagnosis | | 55.26 (0.183) | 54.55 (0.318) | 56.21 (0.817) | 57.74 (1.681) |
| Use cochlear implant | Yes | 0.1% (0.0%) | 0.1% (0.0%) | Masked | - |
| Age started cochlear implant | | 59.98 (1.740) | 65.45 (2.801) | Masked | - |
| Use hearing aid | Yes | 14.3% (0.2%) | 13.3% (0.4%) | 18.2% (1.1%) | 18.9% (2.3%) |
| Age started hearing aid | | 64.54 (0.162) | 64.12 (0.269) | 65.79 (0.587) | 67.17 (1.073) |
| Use assistive listening device | Yes | 0.6% (0.0%) | 0.5% (0.1%) | Masked | Masked |
| Age started assistive device | | 62.29 (0.663) | 63.93 (1.124) | 63.57 (2.994) | 64.86 (2.650) |
| No hearing devices | Yes | 85.2% (0.2%) | 86.3% (0.4%) | 81.5% (1.2%) | 80.5% (2.3%) |
| Ménière's disease | Yes | 2.2% (0.1%) | 2.1% (0.2%) | 2.7% (0.5%) | Masked |
| Vertigo | Yes | 38.8% (0.3%) | 39.0% (0.5%) | 40.4% (1.5%) | 44.3% (2.9%) |
| Tinnitus | Yes | 37.1% (0.3%) | 37.9% (0.5%) | 40.0% (1.5%) | 34.0% (2.8%) |
| Tinnitus severity (0-10) | | 2.54 (0.025) | 2.57 (0.042) | 2.35 (0.107) | 2.33 (0.230) |

*Note.* Categorical measures presented as % (SE), continuous as Mean (SEM). E4+ indicates APOE E4 carriers, E4- non-carriers. Indented items are contingent follow-ups. Statistics with n<5 masked for privacy; dashes indicate not applicable; PD=Parkinson's disease; SRT=speech reception threshold..

### 3 References

23andMe, Inc. (2023). *23andMe® Personal Genome Service® (PGS) Package Insert.* https://permalinks.23andme.com/pdf/package_insert_v5.pdf
